# Effectiveness of prolotherapy vs local anesthetic infiltration guided by ultrasound in the treatment of shoulder pain syndrome

**DOI:** 10.1101/2020.12.22.20248223

**Authors:** Juan A. Lira-Lucio, Guillermo Ochoa-Gaítan, Lizeth Hernández-Escobar, Christian I. Padilla-Rivera, Berenice C. Hernández Porras, Ángel M. Juarez-Lemus, Jose Guillermo Ochoa-Millan, Roberto J. Jimenez-Contreras, Enrique Roldán-Rodríguez

## Abstract

Chronic Shoulder Pain (CSP) is a health problem that affects almost 67% of the general population. Almost a third of patients with acute shoulder pain syndrome don’t respond to initial therapy with analgesics and need interventional therapy. Corticosteroid injection is the standard therapy. Prolotherapy has been demonstrated to be effective in other chronic pain syndromes, but not in CSP. The aim of this study was to determine the effectiveness of prolotherapy compared to local anesthetic injection in the treatment of chronic shoulder pain

**METHODS:** Retrospective and comparative study of 77 patients from the National Institute of Oncology in Mexico City who received treatment for Chronic Shoulder Pain guided by ultrasound between 2017-2019. 57 patients were kept in the study for further analysis. 39 received infiltration with corticosteroids and 17 prolotherapy. Effectiveness of therapies was determined based on the decrease in VAS score in next follow-up session. Statistical analysis were performed with SPSS and RStudio Software.

**RESULTS:** 51% of patients with Chronic Shoulder Pain were unemployed. 84% of the patients needed 3 different types of analgesics before they received ultrasound guided local treatment. Prolotherapy was as efficient as local anesthetic injection, no matter basal pain severity or underlying shoulder diagnosis, despite prolotherapy being more used as treatment for Rotator Cuff Tendinopathy.

**CONCLUSIONS:** Prolotherapy and corticosteroid injection guided by ultrasound have the same efficacy in pain relief for chronic shoulder pain in oncologic patients.

## 1. Introduction

Chronic Shoulder Pain (CSP) is a health problem that affects almost 67% of the general population with high economic and lifestyle burden ^1^. Patients with an oncologic disease have an increased risk of developing shoulder pain after surgical interventions, radiotherapy, and the pathologic features of their underlying disease23. Even with pharmacologic treatment, almost a third part of patients with acute shoulder pain syndrome don’t respond to initial therapy with acetaminophen, non-steroidal anti-inflammatory drugs (NSAIDs) or muscle relaxants and will develop chronic shoulder pain.^4^ If initial therapy, such as NSAIDs, rest, and physical rehabilitation, fail to relieve pain and improve function, the second line of treatment may non-invasive treatment as local anesthetic injection or prolotherapy ^5^.

Local corticosteroid (CS) infiltration is the second most common non-surgical therapy used to treat CSP, after the use of multiple analgesics. Almost 11% of patients with shoulder pain receive CS local infiltration in primary care ^6^. There is uncertainty about CS injection efficacy as therapy of shoulder pain after recent evidence that demonstrate a small and transient pain relief without additional benefit to other therapies. Anesthetic infiltration and physical therapy show equal long-term results for function, range of motion and patient-perceived improvement ^78^. Additionally, CS injections display multiple adverse effects, such as rotator cuff tendon degeneration, exacerbation of neuropathic pain and delay in tissue repair and tendon necrosis, possibly secondary to an increase of oxidative stress by an increase of glutamate receptor NMDAR1 that promote apoptosis after injection of CS9.

Prolotherapy (PT) is a non-surgical technique for the treatment of chronic painful musculoskeletal conditions. It’s based in the infiltration of local tissues with irritating agents to promote fibrous repair in tissues like tendons, joints, or damaged ligaments ^10^. The most common irritating agents used are hyaluronic acid, hypertonic dextrose, zinc, growth hormone, and autologous cells such as platelet-rich plasma11. PT has won field in recent years as treatment elected by patients, physicians and researchers. In figure 1 most common words used in PubMed publication about PT are represented. PT has demonstrated beneficial effects in function, pain relief, and quality of life in patients with osteoarthritis, plantar fasciitis and adhesive capsulitis; with high treatment adherence and patient satisfaction ^12 13^. Bertrand et al. demonstrated prolotherapy efficacy vs placebo in Painful Rotator Cuff Tendinopathy treatment with pain improvement and a higher patient satisfaction.^14^

**Figure 1.**
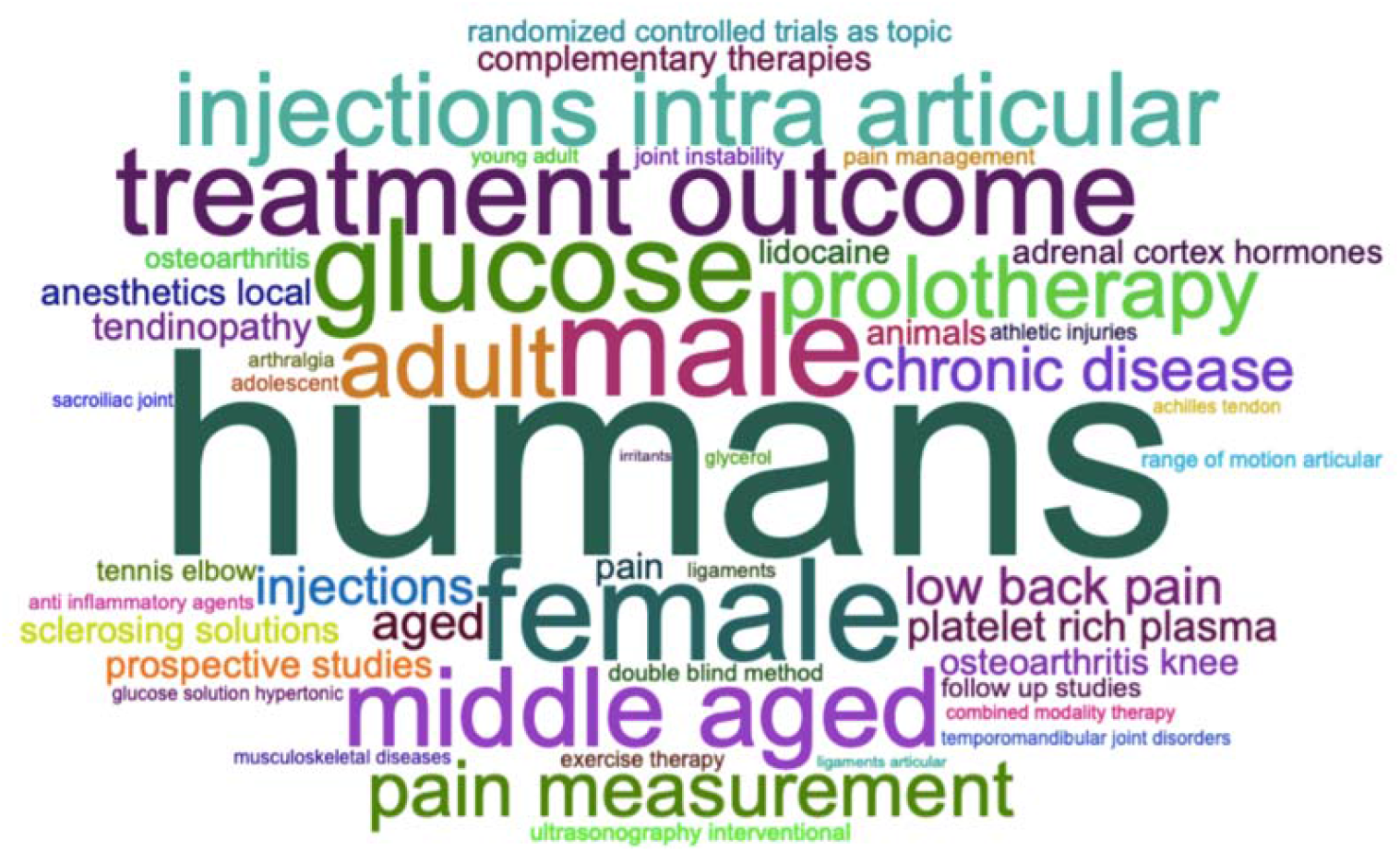
Word Map of Prolotherapy research in PubMed. The size of the words represent the frequency that this words have been used in PL research.

An ultrasonography (US) approach has recently been added to the management of interventional shoulder pain. Therapy guided by ultrasonography (US) is an economic and fast tool to improve previously needle blinded procedures. It has the advantage of being less traumatic to tissues, because it allows puncture to the exact site of infiltration, making this therapy more accurate ^15,12^. PT has not been studied previously in should pain syndrome.

Both techniques have been widely used in pain control with local anesthetic infiltration, but the superiority for pain relief in these patients has not been compared. The aim of this study is to compare the efficacy of prolotherapy versus local anesthetic infiltration for pain relief guided by ultrasonography in patients with chronic shoulder pain.

## 2. Materials and methods

### 2.1 Study Design

Retrospective and comparative study of 77 patients recorded from the National Institute of Oncology in México City with diagnosis of CSP who received US guided interventional pain management between January 2017 and December 2019. Demographic data were collected from medical records and captured in a database.

### 2.2 Studied Population

Inclusion criteria were >18 years and CSP secondary to capsulitis, rotator cuff syndrome, or impingement syndrome. Exclusion criteria were incomplete medical records, patients with a primary tumor in the shoulder, combined technique, or history of previously infiltration. Other pathologies were excluded because low frequency (n=<2) with low statistical representation. The studied population was divided into patients who received nervous infiltration with local anesthetic, and those who were treated with prolotherapy. Mild efficacy of treatment was considered as relief of less than 30% of basal AVS score previous the intervention, moderate efficacy a reduction of 30-50% and, strong efficacy a decrease of >50% from basal AVS score. The present study has been carried out under the Helsinki principles with number of approbation INCAN 2019/0140 by the local bioethics committee.

### 2.3 Statistical analysis

Statistical analysis was performed under IBM^®^, SPSS^®^ software Version 25, WordMap was created in RStudio V 2.3.1 for MacOs Catalina Version 10.15.5 with Bibliometrix package ^16 17 18^. Data distribution was calculated with Shapiro-Wilk test. Data is represented with median and standard deviations (SD). Median differences were determined by T student test; for categorical data X2 test and Fisher Exact Test were used as required. Statistical significance was considered with a p-value <0.05.

## 3. Results

### 3.1 Demographic patient characteristics

77 patients received pain treatment guided by US between January 2017 and December 2019. 57 patients were selected according to inclusion and exclusion criteria. 39 were assigned to the local anesthetic with corticosteroids group (CS) and 17 patients to the prolotherapy group (PT). Demographic features are represented in table 1. There were no differences in age, PT group was 60 (±12.4) vs 60 (±14.06) in LA (p>0.05), 78% of PT patients were women vs 82% in LA but there was not statistical significance. 51% of the patients in both groups were unemployed when the first session of treatment was received. Prolotherapy treatment was more used when the left shoulder was affected. CS therapy was also used more when rotator cuff was affected (90% vs 83%, p<0.0.001). Tumors in the stomach and kidneys were more frequent when prolotherapy was used. 84% of patients in both groups needed treatment with three different families of analgesics before US-guided pain treatment without differences between groups. There was no difference in basal severity of pain between the groups.

**Table I.**
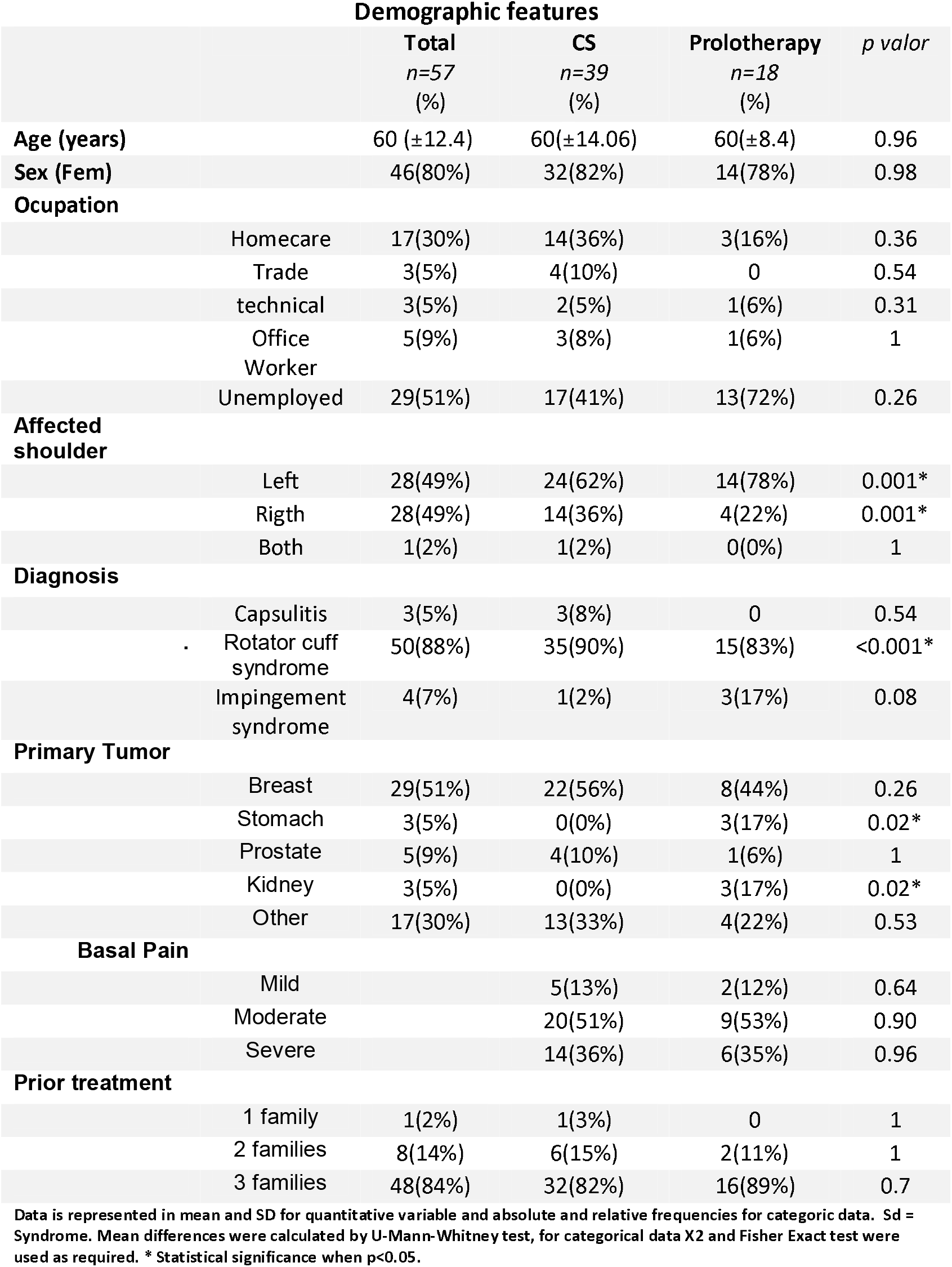

### 3.2 Effectiveness in pain control

Pain control was categorized as mild, moderate, or strong. First, we wanted to know if patient choice of the therapy was associated to basal pain severity, but we did not find any difference (**fig 2**). We then evaluated if pain control was dependent on the initial pain, **table 2**. PT demonstrated to be equal effective as CS to control pain, no matter the basal pain severity. Both groups had good results decreasing chronic pain in patients; 0% in both groups reported mild or no control of pain, 30% in CS had mild control vs 47% in PT group, and respectively 27% vs 56% reported strong control, without statistical difference between the groups (p=0.16). **(Fig 3**).

**TabLE II.**
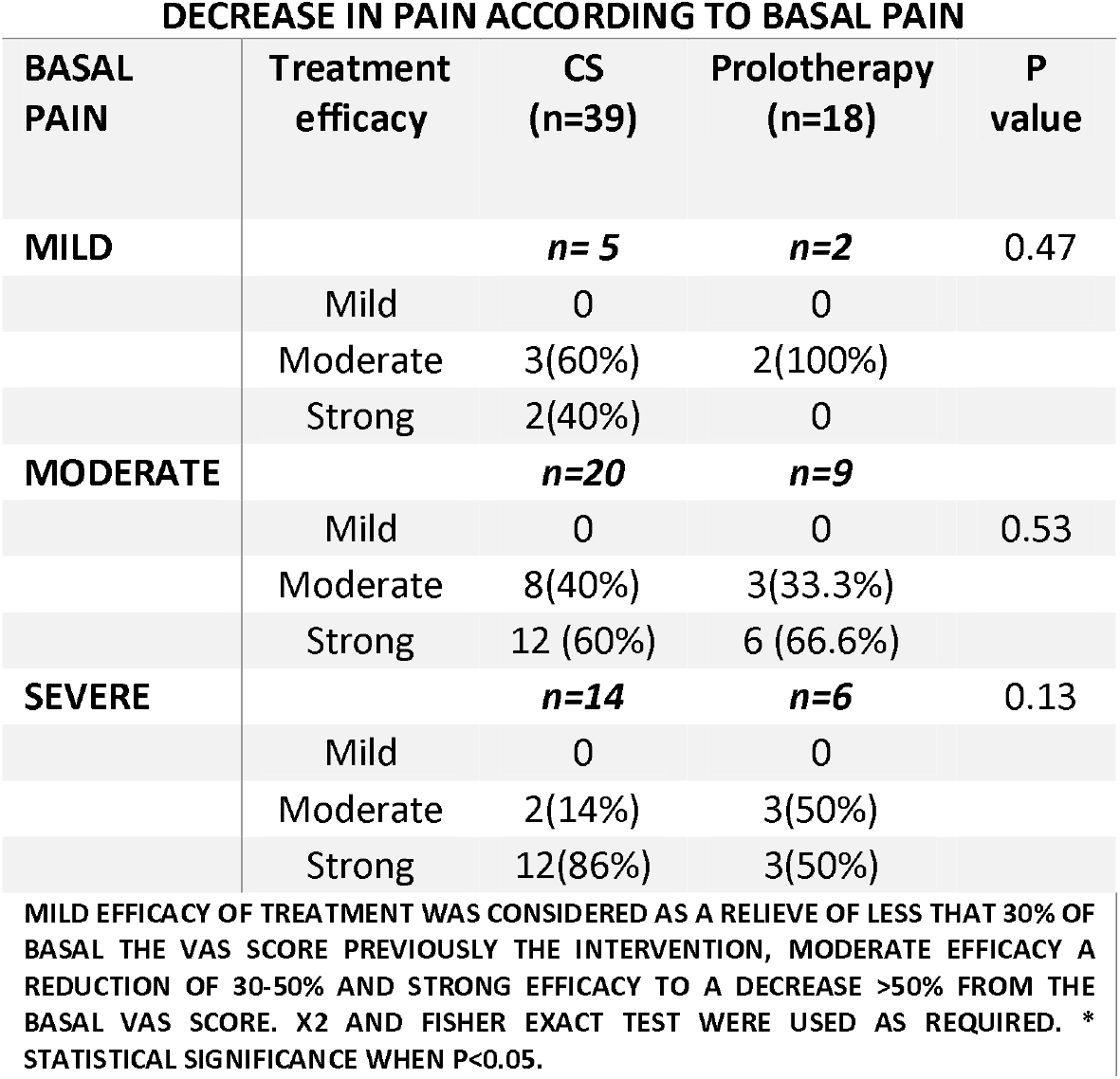
II DECREASE IN PAIN ACCORDING TO BASAL PAIN

| **BASAL PAIN** | **Treatment efficacy** | **CS (n=39)** | **Prolotherapy (n=18)** | **P value** |
| --- | --- | --- | --- | --- |
| **MILD** |  | **n= 5** | **n=2** | **0.47** |
|  | Mild | 0 | 0 |  |
|  | Moderate | 3(60%) | 2(100%) |  |
|  | Strong | 2(40%) | 0 |  |
| **MODERATE** |  | **n=20** | **n=9** |  |
|  | Mild | 0 | 0 | **0.53** |
|  | Moderate | 8(40%) | 3(33.3%) |  |
|  | Strong | 12 (60%) | 6 (66.6%) |  |
| **SEVERE** |  | **n=14** | **n=6** | **0.13** |
|  | Mild | 0 | 0 |  |
|  | Moderate | 2(14%) | 3(50%) |  |
|  | Strong | 12(86%) | 3(50%) |  |

MILD EFFICACY OF TREATMENT WAS CONSIDERED AS A RELIEVE OF LESS THAT 30% OF BASAL THE VAS SCORE PREVIOUSLY THE INTERVENTION, MODERATE EFFICACY A REDUCTION OF 30-50% AND STRONG EFFICACY TO A DECREASE >50% FROM THE BASAL VAS SCORE. X2 AND FISHER EXACT TEST WERE USED AS REQUIRED. \* STATISTICAL SIGNIFICANCE WHEN P<0.05.

**Figure 2.**
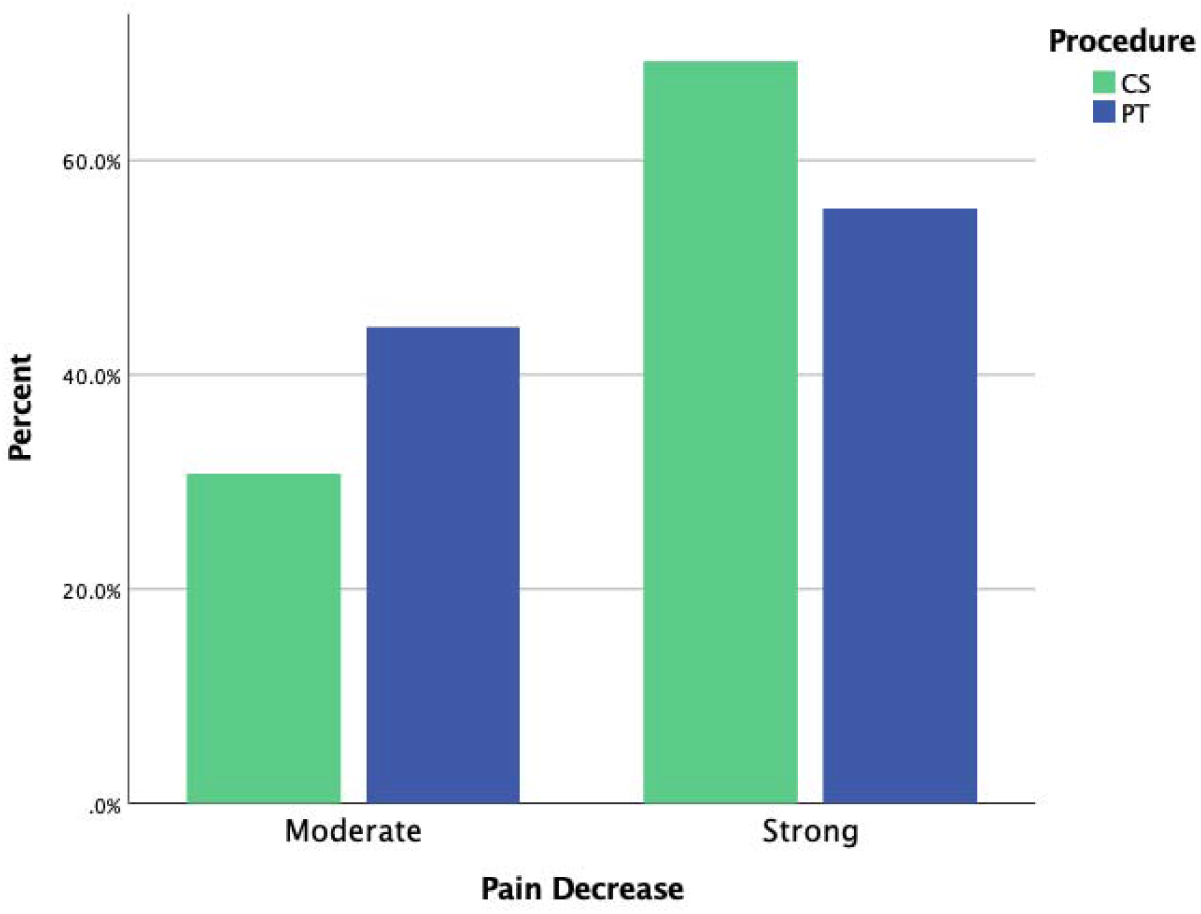
Pain decrease by US guided procedure

**Figure 3.**
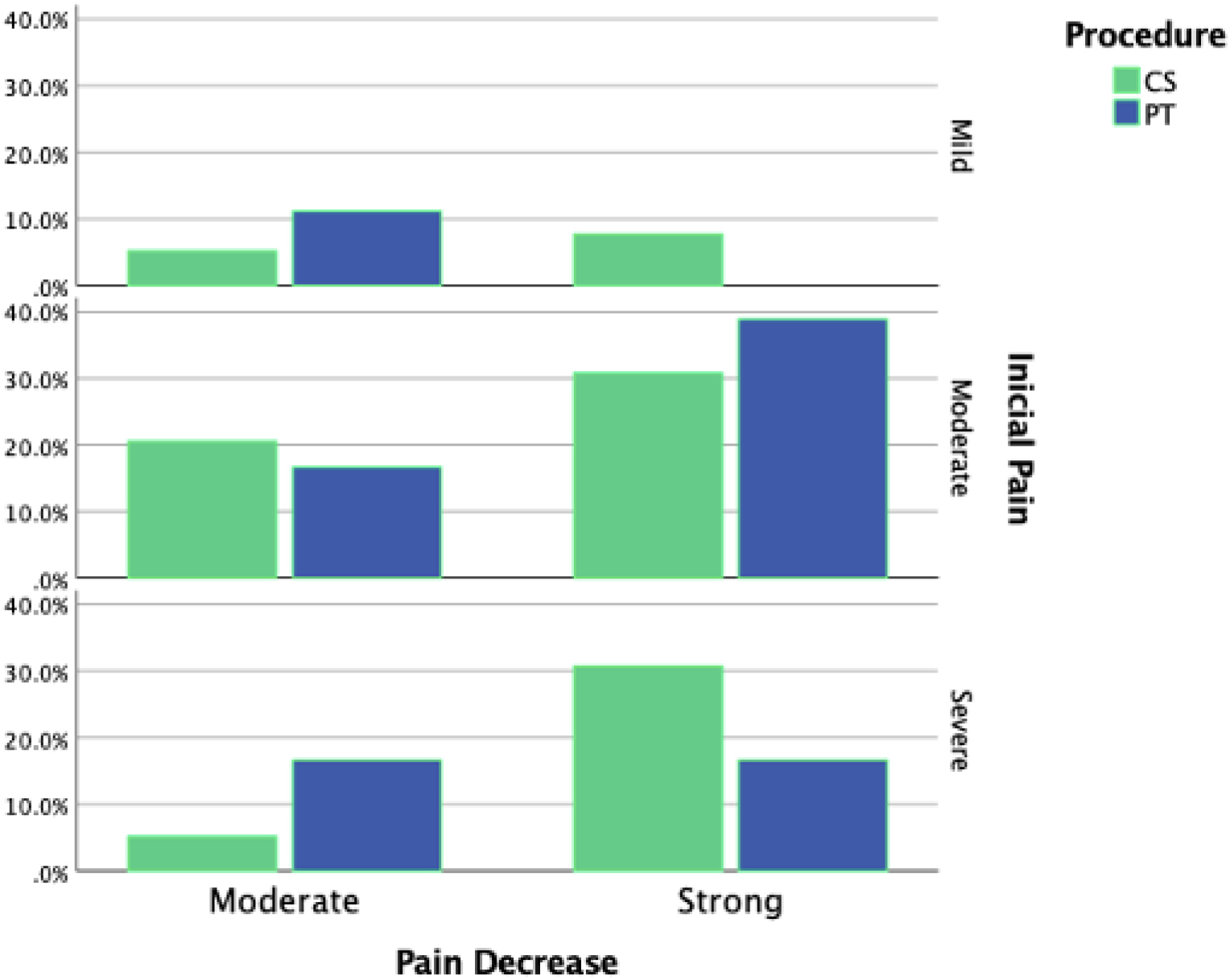
Pain decrease based on initial severity.

CS infiltration was more used in the treatment of rotator cuff syndrome (RCS) than prolotherapy. We evaluated if there is a difference in pain decrease based on the primary diagnosis of CSP. 35 patients in the CS group have RCS vs 15 in the PT group. PT group was equally effective as CS in pain control, with respectively 34% of patients with moderate control vs 45%, and 66% vs 55% with strong relief of pain (p<0.21). This control was not dependent of the number of different analgesics required after infiltration procedures, none of the groups needed 3 families of analgesics. 83% of CS group needed 2 classes of analgesics, vs 57% PL. In the population with strong control, 56% and 50% required 2 classes of analgesics, respectively. None of the participants in this study presented adverse events.

## 3. Discussion

To our knowledge, this is the first study to evaluate the efficacy of CS vs PT for shoulder pain management. PT has demonstrated to be safe and effective in chronic pain therapy in many musculoskeletal pathologies, but just a few studies have investigated PT in CSP, even though it is the third most common musculoskeletal disorder in general practice6. As in other studies reported previously, in this age group the main cause of CSP was Rotator Cuff Syndrome, with a frequency of 88% in the population of this study. This indicates that even though oncologic patients have an increased risk of CSP, the main cause is still Rotator Cuff Syndrome.^15 6^There is no clear indication of prolotherapy for pain treatment in shoulder pathologies, but in this study, we have demonstrated that it is equally effective as CS injections for treatment of Capsulitis, Rotator Cuff Syndrome and adhesive capsulitis. Further studies are needed to evaluate each of the procedures separately in CSP.

Interestingly, almost 50% of patients in both groups were unemployed when they received local therapy. This agrees with other studies about the potential functional restriction that CSP leads to in patients, and the importance of a therapy that improves these functional restrictions. ^19^Ultrasound guided pain management has been studied widely. This is a useful equipment for shoulder pain management ^15^. Raeissadat et al. demonstrated in a prospective study that CS and PT in patients with plantar fasciitis has the same efficacy in pain relief after a 24-weeks follow up. ^12^ In this study, we have also demonstrated that the use of PT has the same efficacy as LA infiltration for chronic shoulder pain. We also found that pain relief does not depend on basal pain severity. The mechanism of PT effectiveness is based on the induction of anti-inflammatory reactions by irritant agents that enhance tissue healing. ^11^

We are aware of the limitations of this study, as a retrospective study where we lack control of some variables of interest. However, to our knowledge this is the first study to evaluate the efficacy of PT vs CS for CSP. CS therapy requires multiples sessions, in this study PT and LA infiltration demonstrated to be safe when guided by US.

CS and PT therapies have limitations in shoulder pain treatment. In randomized controlled trial, Kesikburun et al demonstrate that PT is not a therapy that should be used alone, its effectiveness is dependent of physical rehabilitation ^20^. In this study, we have evaluated the use of PT and LA nervous infiltration separately. This study allows us to place PT as a safe technique when it is guided by US and enables future blinded studies in shoulder pain. PT is a promising technique, and it has demonstrated that its benefits are not limited to pain control, but also improvement of functionality and mobility.

## Conclusions

Most patients with CSP needs to receive pharmacologic treatment before receiving an interventional management. LA and PT have demonstrated to relieve pain in short term in these patients, regardless of the severity of basal pain. Prolotherapy is a safe and minimally invasive technique with high adhesion to treatment for pain control in CSP patients. It has the same effectiveness in pain relive as CS nervous injection in oncologic patients, no matter the basal severity of pain. Further prospective, blinded and randomized studies with covariates as functional improvement are needed to prove PT long term benefits, but this study demonstrate that it is a promising treatment.

### WHAT IS KNOWN

- Patients with painful man syndrome need multiple therapies to have pain relief
- Use of corticosteroid infiltration is therapy with few acute adverse events when performed by ultrasound.

### WHAT IS NEW

- Prolotherapy is as effective as corticosteroid infiltration in pain relief of patients with painful shoulder syndrome.
- When ultrasound guided prolotherapy is a safe therapy for handling painful shoulde syndrome.

## Data Availability

The datasets generated during and/or analysed during the current study are available from the corresponding author on reasonable request

## Conflicts of interest

The authors certify that there is no conflict of interest with any financial organization regarding the material discussed in the manuscript.

## Funding

The authors report no involvement in the research by the sponsor that could have influenced the outcome of this work.

## Author Contributions

ER-R, Data collection and curation; JAL-L and GO-G, Formal analysis; JAL- and LH-A, Investigation; ER-R and AJ-L Methodology; JAL-L, COG-G, ER-R, Project administration; JAL-L, Software; GO-G and ER-R, Supervision; JAL-L, GO-G, LH-A, AJ-L ER-R, Validation; JAL-L and LH-A, Visualization; JAL-L and ER-R, Writing—original draft preparation; GO-G, LH-A – review & editing. All authors have read and agreed to the published version of the manuscript.

…

## Notes

### Competing Interest Statement

The authors have declared no competing interest.

### Author Declarations

This is a retrospective study using clinical data from the clinical record of patients from the National Institute of Oncology (INCan)in Mexico City. The protocol has reviewed by Institucional Bioethics Committee "Comité de Ética en Investigación, Instituto Nacional de Cancerología, Ciudad de México" and ethical approval was given in accordance with the Declaration of Helsinki (Minutes of approval: INCAN 2019/0140).

